# INSPECT-SR: a tool for assessing trustworthiness of randomised controlled trials

**DOI:** 10.1101/2025.09.03.25334905

**Authors:** Jack Wilkinson, Calvin Heal, Ella Flemyng, Georgios A. Antoniou, Tony Aburrow, Zarko Alfirevic, Alison Avenell, Virginia Barbour, Vincenzo Berghella, Dorothy V. M. Bishop, Esmée M Bordewijk, Nicholas J. L. Brown, Jana Christopher, Mike Clarke, Darren Dahly, Jane Dennis, Patrick Dicker, Jo Dumville, Helen Frankish, Andrew Grey, Steph Grohmann, Lyle C. Gurrin, Jill A. Hayden, James A.J. Heathers, Kylie E Hunter, Ian Hussey, Lukas Jung, Emily Lam, Toby J. Lasserson, Sarah Lensen, Tianjing Li, Wentao Li, Jianping Liu, Elizabeth Loder, Andreas Lundh, Gideon Meyerowitz-Katz, Ben W. Mol, Florian Naudet, Anna Noel-Storr, Neil E. O’Connell, Lisa Parker, Rita F. Redberg, Barbara K. Redman, Rachel Richardson, Anna Lene Seidler, Kyle Sheldrick, Emma Sydenham, Madelon van Wely, Colby J. Vorland, Rui Wang, Stephanie Weibel, Matthias Wjst, Lisa Bero, Jamie J. Kirkham

## Abstract

The integrity of evidence synthesis is threatened by problematic randomised controlled trials (RCTs). These are RCTs where there are serious concerns about the trustworthiness of the data or findings. This could be due to research misconduct, including fraud, or due to honest critical errors. If these RCTs are not detected, they may be inadvertently included in systematic reviews and guidelines, potentially distorting their results. To address this problem, the INSPECT-SR (INveStigating ProblEmatic Clinical Trials in Systematic Reviews) tool has been developed to assess the trustworthiness of RCTs. This will allow problematic RCTs to be identified and excluded from systematic reviews. This paper describes the development of INSPECT-SR. The tool and an associated guidance document are presented.

## Introduction

In systematic reviews of randomised controlled trials (RCTs) the established steps are to identify all eligible trials, to appraise them using risk of bias tools, and to synthesise the results to reach a conclusion about the effectiveness and harms of an intervention. This paradigm is threatened by the presence of *problematic studies* in the literature. As defined by Cochrane, these are studies “where there are serious questions about the trustworthiness of the data or findings” (1). A study could be problematic due to research misconduct (including plagiarism, falsification, and fabrication of data or methods) or due to critical errors in study conduct that would not be flagged by risk of bias tools. If these studies are not identified as untrustworthy, they may inadvertently be included in evidence synthesis, potentially leading to treatment recommendations that are misleading or even actively harmful (2–4). Because many of these studies are not easily identifiable, it is difficult to assess the prevalence of problematic RCTs, but the available evidence is not encouraging. Carlisle estimated that 26% of RCTs submitted to the journal Anaesthesia were problematic (5). A recent review identified 847 systematic reviews that included one or more retracted RCTs in meta-analysis, and found that excluding these studies changed the size of effect in 16% of meta-analyses, and the direction of effect in just over 8% (6). It may take years for a problematic trial to be recognised and retracted, if it is retracted at all (7, 8), so these figures are likely to underestimate the problem. These delays also mean that systematic reviewers cannot rely on retraction status alone to identify problematic trials.

Risk of bias tools are not designed to identify problems of this nature, nor do they appear to be successful in doing so (9, 10). Bespoke tools for *trustworthiness assessment* are therefore needed. A variety of trustworthiness tools have recently been proposed (11–15), although they differ in terms of their development, content and structure, and the appropriateness of some of the trustworthiness checks that have been included in some of these tools is unclear (16). Given the number of problematic studies that are now being identified, and the growing number of proposed methods for detecting these studies, it was timely to assess and reach international consensus on a set of checks and principles for trustworthiness assessment.

In this article, we present the INSPECT-SR (INveStigating ProblEmatic Clinical Trials in Systematic Reviews) tool. INSPECT-SR implements a set of trustworthiness checks, selected on the basis of empirical evidence, expert consensus methods, and theoretical considerations in the form of a practical tool that will help systematic reviewers and users of primary research to assess whether an RCT is trustworthy.

### Scope of the INSPECT-SR tool

INSPECT-SR has been created to assess trustworthiness of RCTs in order to identify problematic studies. This involves checking various aspects of the trial publication and associated documents in order to reach a conclusion about the veracity of the study’s reported methods and results. The tool does not assess risk of bias, generalisability, or conflicts of interest. The tool does not require that individual participant data (IPD) are available. A tool for assessing trustworthiness using IPD has been developed (15) and INSPECT-IPD, an extension to INSPECT-SR that can be used when IPD are available, is in development (funder ref: NIHR303741). INSPECT-SR can be used on all RCTs, regardless of the area of healthcare. The tool has not been designed as a diagnostic test for fraud, since honest error that is fatal to an RCT’s conclusions cannot generally be ruled out, and it should not be deployed or interpreted as such. Concerns about a trial’s trustworthiness do not constitute an accusation of research misconduct.

### Development of the tool

The development process for the tool has previously been described (17) and included five stages: 1) creation of a comprehensive list of trustworthiness checks using existing literature and a survey of experts, to be evaluated in subsequent stages; 2) application of the checks to 50 Cochrane Reviews to evaluate their feasibility and impact; 3) a Delphi survey to establish which of the checks are supported by expert consensus; 4) a series of online consensus meetings to determine which checks to include in the draft INSPECT-SR tool, and how they should be operationalised; and 5) user testing of the draft tool, with feedback, captured using an online survey and user workshop, to finalise the tool and accompanying guidance document. Stages 1 and 2 have been reported previously (9, 18). Here we report Stages 3 to 5 and present the final tool. Figure 1 shows the number of checks included at each stage of development.

**Figure 1:**
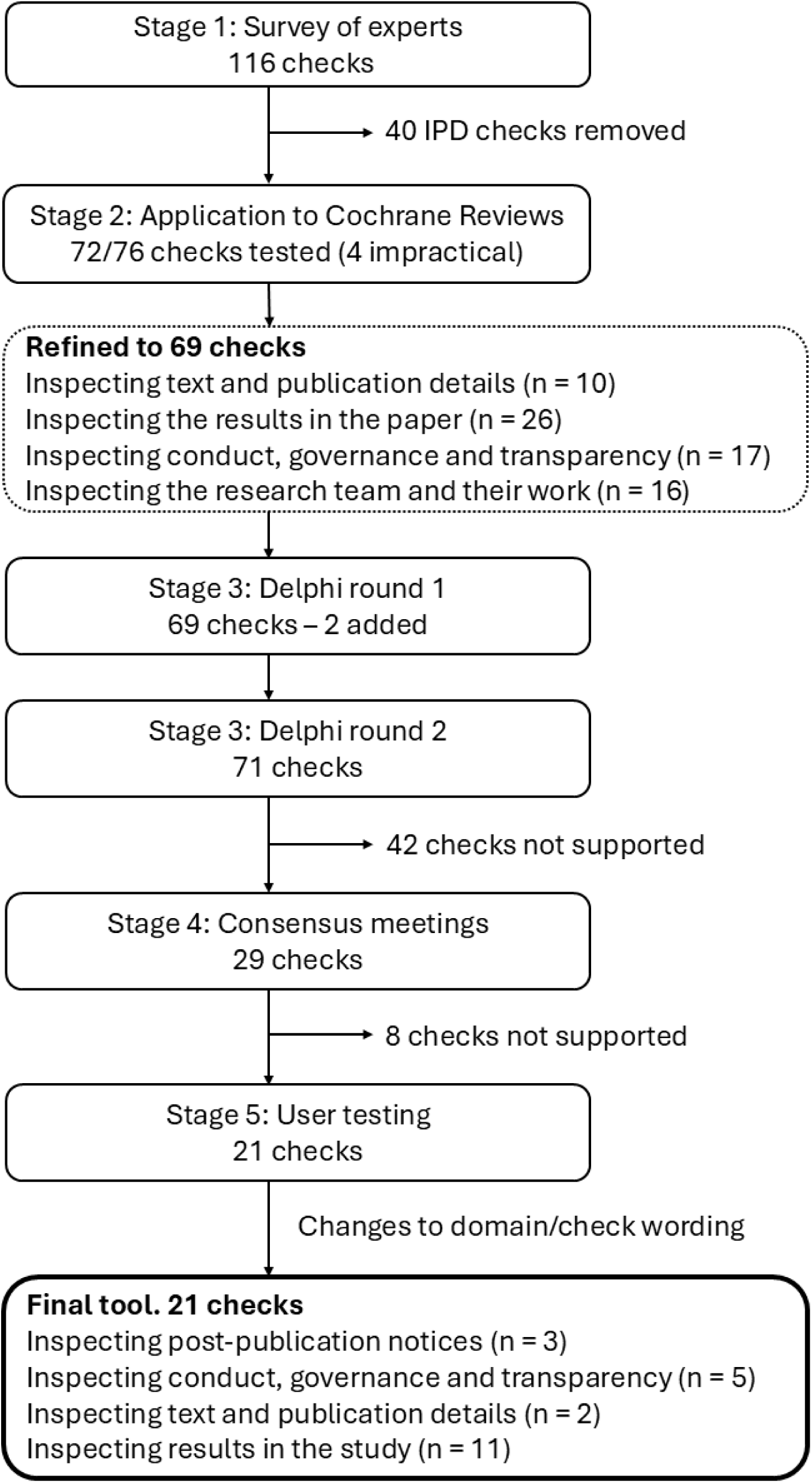
Summary of the development of INSPECT-SR

#### Candidate item list generation

A comprehensive list containing 76 trustworthiness checks was produced using existing evidence (a scoping review (19) and qualitative study (20)) and a new survey of experts (18). The list was refined in consultation with the project expert advisory panel using results from an application of the checks to RCTs in Cochrane Reviews (9). The refined list of 69 checks and associated explanations, arranged in four domains (Figure 1), can be viewed at https://osf.io/esxu7.

#### Recruitment of Delphi participants

Individuals with expertise or experience in assessing potentially problematic studies and potential users of the tool were eligible to participate. Individuals were identified via professional networks of the study team and expert advisory panel, from individuals who had contacted the study team expressing interest, from authors of relevant publications, and via advertisement on social media and academic presentations. Participants were invited by personalised email, containing a unique participation link. Consent was obtained within the survey. Eligible individuals were invited to participate in Round 2 even if they had not participated in Round 1.

#### Delphi survey

The survey was implemented in Qualtrics (Provo, UT) and can be viewed at https://osf.io/9nu23. Participants were asked to score each check 1 to 9 on two scales, relating to usefulness and feasibility. Usefulness was explained with the text “Do you think performing this check would help to identify problematic studies?” Feasibility was explained with the text “Do you think performing this check would be easy for a competent review team to perform when assessing trustworthiness of a study, in terms of required skill, expertise, resources, or time?” A “don’t know” option was also available. Free text boxes were included for comments, or for suggestions for additional checks, which were then included in the Round 2 survey. At Round 2, participants were presented with their own scores from Round 1, and a summary of scores from all participants. Participants were then asked to score each check again in light of the Round 1 scores. A prespecified consensus threshold was applied. Any check that scored 7 or more for usefulness by at least 80% of participants at Round 2 was considered to be supported.

The Delphi was conducted between November 2023 and April 2024. 323 and 333 individuals were invited to Rounds 1 and 2, respectively, with 148 and 134 participants completing each round. Supplementary Table 1 shows characteristics of participants. Twenty-nine checks met the consensus criterion and were then entered into the Stage 4 consensus meetings (Supplementary Table 2).

#### Consensus meetings

For each of four domains, a consensus meeting was held in duplicate (eight total meetings) to accommodate participants in different time zones. Different participants were invited to meetings relating to different domains, to ensure that checks were discussed by people with relevant expertise. Results from Stages 2 and 3 were presented to participants (slides can be viewed at https://osf.io/5k7yf/). Following a short discussion, participants used Mentimeter to vote anonymously on whether the check should be included in the tool. Because each meeting was held in duplicate, there was scope for disagreement between groups. Where this occurred, participants from both meetings were invited to anonymously present arguments for and against the check using a collaborative online tool (Google Jamboard). All participants were subsequently invited to vote again via an online survey, implemented in Qualtrics, and the consensus criterion was applied to determine inclusion.

The meetings were held in June and July 2024. Twenty-one checks were selected for inclusion on the basis of the consensus meetings and subsequent resolution of four disagreements (Supplementary Table 3). A draft tool was created including these 21 checks, which were reorganised and reworded in light of the consensus meeting discussions. One domain was renamed, one was dropped, and a new domain was introduced.

#### User testing of the draft tool

In Stage 5, the draft tool was tested and feedback was gathered from testers. Individuals who appraise RCTs as part of their role were recruited. Individuals were supplied with the draft tool and a summary guidance document, were asked to use these to assess the trustworthiness of one or more RCTs, and to provide feedback via an online survey (available at https://osf.io/y2c5u). Participants were supplied with a participant information sheet and were asked to provide written consent to take part before being supplied with a personalised survey link. Participants were asked whether they would be willing to discuss their feedback at a user workshop (slides available at https://osf.io/truwq).

Survey responses were collected between October 2024 and February 2025. Supplementary Table 4 shows a summary of responses. 148 individuals were invited to provide feedback, and 75 gave written consent to participate. 40 individuals provided feedback via the survey. The median (IQR) number of trials assessed was 3 (1.5 to 4). The median (IQR) time to assess an RCT using the tool was 45 (27 to 74) minutes. 28 (70%) participants said they would choose to use the tool in future work, with 11 (28%) saying they did not know whether or not they would do so. One person said they would not. 37 (93%) said that they thought the judgements they reached using the tool were reasonable. Challenges with implementing several checks were highlighted.

The user feedback workshop was attended by 8 participants on 14th January 2025. Anonymised minutes from the meeting are shown in Supplementary Table 5. It was determined at the workshop that some challenges and user questions could be addressed by expanding the associated guidance document, and by providing a collection of examples for users of the tool. It was also determined that software would be useful to assist in the implementation of some of the checks, and that the guidance document should direct users to this software where available. The feedback was used to make minor changes to the tool (numbering for checks was added, some checks were reworded). In addition, a detailed version of the guidance document was produced, including examples of each check.

### The INSPECT-SR tool

INSPECT-SR guides a reviewer through a series of up to 21 checks organised in four domains 1) *Inspecting post-publication notices* (3 checks), 2) *Inspecting conduct, governance, and transparency* (5 checks), 3) *Inspecting text and figures* (2 checks), 4) *Inspecting results in the study* (11 checks) (Table 1). An answer of “Yes” in response to a check indicates a potential problem. After completing the checks in a domain, the tool prompts the reviewer to make a judgement about the trustworthiness of an RCT in relation to that domain (“no concerns”, “some concerns”, or “serious concerns”), and to make an overall judgement about the trustworthiness of the study on the basis of the domain-level judgements (Figure 2). A detailed guidance document, including examples of each check, and an editable Microsoft Word template are provided at https://osf.io/b74wj.

**Figure 2:**
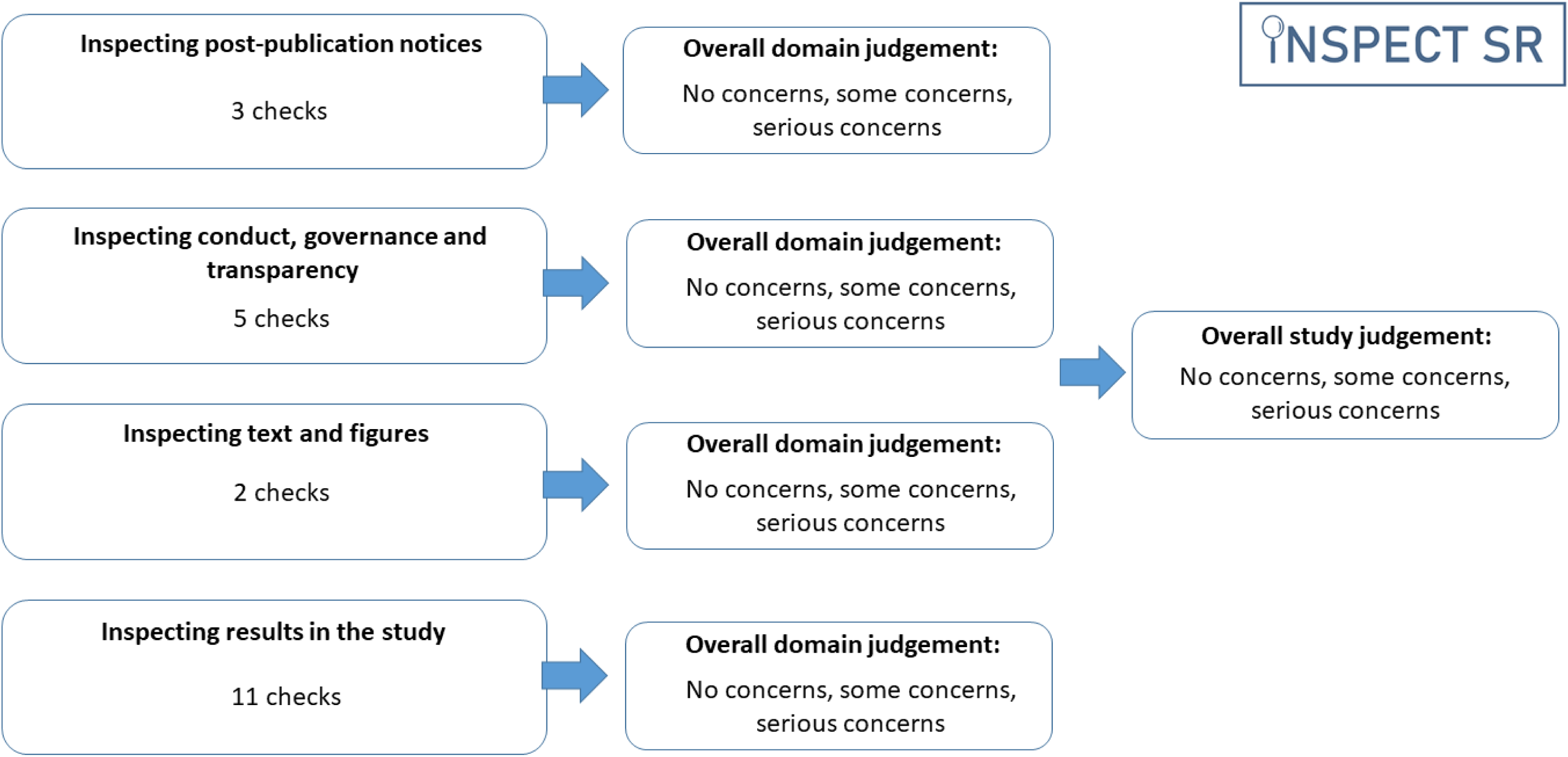
Structure of the INSPECT-SR tool

**Table 1:** INSPECT-SR.

| Domain and check | Response |
| --- | --- |
| **Inspecting post-publication notices** |  |
| 1.1. Does the study have an associated retraction? | Yes, No, Unclear or Not applicable |
| 1.2. Does the study have an associated expression of concern or other relevant post publication notice? | Yes, No, Unclear or Not applicable |
| 1.3. Do other studies by the research team highlight causes for concern (associated retractions, expressions of concern, relevant post-publication notices?) | Yes, No, Unclear or Not applicable |
| **Overall domain judgement** | **No concerns, some concerns, serious concerns** |
| **Inspecting conduct, governance, and transparency** |  |
| 2.1. Are there concerns relating to ethical approval? | Yes, No, Unclear or Not applicable |
| 2.2. Are there concerns relating to the timing or absence of study registration? | Yes, No, Unclear or Not applicable |
| 2.3. Are there important inconsistencies between the publication and the registration documents? | Yes, No, Unclear or Not applicable |
| 2.4. Is the recruitment of participants implausible? | Yes, No, Unclear or Not applicable |
| 2.5. Are the reported methods implausible considering the reported resources? | Yes, No, Unclear or Not applicable |
| **Overall domain judgement** | **No concerns, some concerns, serious concerns** |
| **Inspecting text and publication details** |  |
| 3.1. Are there concerns relating to duplicated content, such as text or tables, or text that is incompatible with the study? | Yes, No, Unclear or Not applicable |
| 3.2. Is there evidence of manipulation or duplication of figures? | Yes, No, Unclear or Not applicable |
| **Overall domain judgement** | **No concerns, some concerns, serious concerns** |
| **Inspecting results in the study** |  |
| 4.1. Are there any unexplained discrepancies between reported data and participant eligibility criteria? | Yes, No, Unclear or Not applicable |
| 4.2. Are numbers of participants allocated to each group implausible given the allocation method? | Yes, No, Unclear or Not applicable |
| 4.3. Are any baseline data implausible? | Yes, No, Unclear or Not applicable |
| 4.4. Are there any discrepancies between results reported in figures, tables, and text? | Yes, No, Unclear or Not applicable |
| 4.5. Are the numbers of participants lost to follow-up implausible? | Yes, No, Unclear or Not applicable |
| 4.6. Are there any unexplained inconsistencies in the numbers of participants? | Yes, No, Unclear or Not applicable |
| 4.7. Are any outcome data, including estimated treatment effects, implausible? | Yes, No, Unclear or Not applicable |
| 4.8. Are the means and variances of integer data impossible? | Yes, No, Unclear or Not applicable |
| 4.9. Are there errors in statistical results? | Yes, No, Unclear or Not applicable |
| 4.10. Are any other contradictions implied by the data? | Yes, No, Unclear or Not applicable |
| 4.11. Are there inconsistencies in descriptions of methods and results across publications describing the study? | Yes, No, Unclear or Not applicable |
| **Overall domain judgement** | **No concerns, some concerns, serious concerns** |
| **Overall study judgement** | **No concerns, some concerns, serious concerns** |

### Using INSPECT-SR

Users should consult the detailed guidance document (https://osf.io/b74wj) before using INSPECT-SR. An overview of key points is provided here.

#### Using check responses to arrive at an overall judgement

The guidance document includes instructions and examples for each check in the tool. The tool does not use a prescriptive algorithm to derive domain-level judgements from check responses. For example, we have not specified a threshold corresponding to a number of “Yes” responses to checks required to arrive at a domain-level judgement of “serious concerns”. Rather, the checks are intended to help the reviewer to reach a domain-level judgement about trustworthiness, and to articulate the rationale for the judgement. An answer of “Yes” to an individual check should not automatically trigger a judgement of “serious concerns”, although in some circumstances the problem highlighted by a check may provide a sufficient basis for doing so. For example, if the answer to check 1.1 *- Does the study have an associated retraction?* is “Yes” then this would usually be sufficient grounds to arrive at a domain-level judgement of “serious concerns”. In principle, a reviewer may decide that they have sufficient evidence to arrive at a domain-level judgement of “serious concerns” at any point during the assessment. If this happens, the reviewer may stop the assessment, assigning “serious concerns” to the study overall.

The overall study-level judgement should typically be at least as severe as the most severe domain-level judgement. This means that the study-level judgement should be at least “some concerns” if one or more domain-level judgements are “some concerns” and no domain-level judgement is “serious concerns”, and should be “serious concerns” if any domain-level judgement is “serious concerns”. A reviewer may also consider a study-level judgement of “serious concerns” to be appropriate on the basis of “some concerns” for several domains. In the event that “serious concerns” are identified, the reviewer should reflect on the rationale for the judgement to ensure that it is warranted. Regardless of the overall judgement, the reasons for the domain and study-level judgements should be clearly reported. The editable template (https://osf.io/b74wj) includes space to add free-text comments.

The domains and checks in INSPECT-SR have been deliberately arranged to support an efficient assessment process, with relatively straightforward but definitive checks, such as checks for post-publication notices, appearing early in the list. However, reviewers may assess domains (or checks within domains) in a different order if they prefer to do so.

#### Incorporating INSPECT-SR into the systematic review process

In the context of a systematic review, we recommend that INSPECT-SR is applied to assess trustworthiness of all eligible RCTs. We advise that INSPECT-SR should be used before risk of bias assessment and data extraction, to avoid wasting time assessing the risk of bias of, and extracting data from, problematic trials. RCTs with “serious concerns” should be excluded from the systematic review. RCTs with “some concerns” should be subjected to sensitivity analysis (for example, by comparing an analysis restricted to RCTs with “no concerns” to one that also includes RCTs with “some concerns”).

We recommend that INSPECT-SR be applied independently by two reviewers, who should then confer to reach consensus in relation to an assessment. It is likely to be beneficial to include at least one reviewer with content expertise as part of the assessment. While methodological expertise is also likely to be useful, use of the tool does not require advanced statistical knowledge. We recommend contacting study authors to attempt to resolve uncertainties. Templates for reporting trustworthiness concerns to journals are available from Cochrane (https://www.cochranelibrary.com/cdsr/editorial-policies/problematic-studies-implementation-guidance#7-1).

## Discussion

INSPECT-SR provides a rigorously developed, systematic and transparent method for identifying problematic RCTs in systematic reviews. It includes a series of checks that have been selected on the basis of empirical evidence and expert consensus. For a majority of reviewers, methods for the assessment of trustworthiness issues of this kind will be unfamiliar. This introduces important considerations for implementation. First, there is a need to have clear guidance to limit misapplication and misinterpretation of the checks contained in the tool. We have developed an accompanying guidance document, accessible at https://osf.io/b74wj, for this purpose. The guidance has been made available in an updatable format, as we anticipate that it will evolve continually in response to user feedback, evaluation of the tool, and developments in software and trustworthiness research. We will also add further training materials to this location.

A second consideration is that there will be a learning curve associated with the use of INSPECT-SR, and it may take longer for novice users to apply the tool. The median time per RCT was reported as 45 minutes in user testing, which is not unreasonable given that these were first-time users with limited guidance to facilitate their assessments. However, there was considerable variation in this assessment time. This appears to be explained by several factors. First, the guidance prompts users to stop the assessment if they judge there to be “serious concerns” at any point, meaning that some assessment times will be very short. Another factor is that some testers opted to review all material related to a RCT - for example, carefully checking results in supplementary materials for errors, or reviewing all versions of the clinical trial registration record. While this will increase the chance of identifying problems, it might not be practical for many reviewers. A pragmatic approach might, for example, involve testing a selection of results for errors, rather than every testable result in the paper. AI-based tools could be useful in this regard in the future, if developed to acceptable standards (21). The use of large language models (LLMs) to expedite systematic review processes is an area of active research (22, 23) and this includes work exploring use of LLMs to assist with INSPECT-SR [UKRI Metascience research grant (OPP569) awarded to Avenell].

Systematic reviewers and guideline developers have a responsibility to identify problematic studies because the trustworthiness of evidence synthesis depends on the evidence it includes. Recent examples highlight the issue. A National Institute for Health and Care Excellence (NICE) recommendation on fetal pillow (Cooper-Surgical) has been reversed following the retraction of an RCT of the device (24). It is possible that a trustworthiness assessment could have prevented inclusion of the trial in the recommendation ahead of its retraction, as it appears to contain statistical anomalies, including in the abstract. During the COVID-19 pandemic, systematic reviews for ivermectin suggested an impressive benefit of the drug for reducing mortality on the basis of potentially problematic trials (25), while subsequent high-quality evidence appears to show modest or no benefit (26, 27). Examples of potentially problematic trials threatening the integrity of systematic reviews can be found across a diverse range of clinical areas (3, 10, 28). The cost of doing nothing at all could be severe.

There have historically been no widely agreed standards of trustworthiness assessment (3), leading to ad-hoc, obscure, and potentially inequitable assessments. INSPECT-SR attempts to address this gap by providing standardised criteria for transparent trustworthiness assessment, which have the backing of a large, international, expert consensus. Systematic reviewers should share the methods that they use for identifying problematic studies, as well as the results of their investigations. INSPECT-SR will enable the equitable application of a systematic method and, by providing a structured format for transparently articulating concerns, will minimise duplication of effort to identify problematic studies. Nonetheless, ongoing evaluation of INSPECT-SR will be important to identify areas of misuse and misinterpretation, to identify training needs, and to identify examples of good practice to use as exemplars. Findings from this work will be used to improve the INSPECT-SR guidance document. In this regard, we would caution against attempts to assess “diagnostic accuracy” of INSPECT-SR, for example, by comparing judgements obtained using the tool to retraction status of articles, if that is based on the assumption that this represents an objective gold standard. In reality, whether or not an article is retracted is a subjective decision that is to some degree arbitrary, such that identification of trustworthiness concerns in a non-retracted article might not reflect a failure of the tool so much as a failure of pre or post-publication editorial processes (16).

While several other trustworthiness tools have been proposed, there are important differences with INSPECT-SR in terms of development, scope, content, and form. For example, recently published trustworthiness guidelines from a collaboration of OBGYN Editors include consideration of adherence to reporting standards and outcome reporting bias (29), which are not within the scope of INSPECT-SR. REAPPRAISED is another tool that includes items that are out of scope of INSPECT-SR, such as consideration of appropriateness of statistical methods and content relating to animal research (30). Items that were in scope in REAPPRAISED were considered for inclusion in INSPECT-SR (18), and many checks eventually included in INSPECT-SR have predecessors in the earlier tool. The INSPECT-SR guidance cautions against some of the checks included in other tools or published guidance, such as applying a penalty for trials that under-recruit.

Routine trustworthiness assessment may act as a deterrent to the production of problematic studies. Excluding problematic studies from systematic reviews has the potential to improve the scientific literature by reducing citations of problematic studies and their use by researchers and policy makers (31, 32). Nonetheless, an important question for developers of trustworthiness tools is whether the tools could be used by fraudsters to produce more convincing forgeries. We expect that many fabricators have produced and published fake research on the reasonable assumption that no one would notice, or even consider the possibility of, the fakery and, on balance, we expect that INSPECT-SR is likely to detect and deter many more fake RCTs than it creates. It is also unclear to what extent efforts to circumvent INSPECT-SR, either by manual fabrication or using LLMs, will be successful. Both of these areas should be a focus of future research.

INSPECT-SR has been developed primarily for assessment of health-related RCTs in a systematic review context. We encourage systematic review producers to adopt INSPECT-SR. We also encourage publishers to support adoption of INSPECT-SR by recommending it in instructions to authors of systematic reviews. Updates to the PRISMA (Preferred Reporting Items for Systematic reviews and Meta-Analyses) statement should include guidance on reporting methods and results of trustworthiness assessment (33). Funders and commissioners of systematic reviews should recognise the value of routine trustworthiness assessment, and should recognise that it may be necessary to increase resources and timelines to support this activity.

Future work will develop versions of the tool for other study designs, including non-randomised studies of interventions. Variations of INSPECT-SR may facilitate identification of problematic studies for some other responsible parties. For example, a version of the tool for use by journal editors in editorial assessment, INSPECT-JR, is planned. While systematic reviewers can draw attention to problematic studies and use INSPECT-SR to improve the trustworthiness of systematic reviews, they cannot prevent publication of problematic studies. As noted previously, everyone involved in the scientific publication pipeline – researchers, journals, publishers, and funders – needs to work together to reduce publication of problematic studies (34).

## Data Availability

Stage 1 data available at https://osf.io/6pmx5/files/osfstorage.
Stage 2 data available at https://osf.io/9pyw2/files/osfstorage.
Stage 3 data available on request to the authors.
Stage 4 data included in the manuscript.
Stage 5 data available on request to the authors.

## Ethics statements

The University of Manchester ethics decision tool was used on September 30, 2022 with respect to Stages 1 to 4. Ethical approval was not required for this study because it involved appraisal of published research, surveying professionals about their area of expertise, and no collection of personal information or sensitive or confidential information. An ethical exemption letter was obtained from the University of Manchester Research Ethics Team with respect to Stage 5 (testing and feedback) on September 25, 2024 [Ref: 2024-21418-37434].

## Declarations

JW, CH, GAA, LB, JJK declare funding from NIHR (NIHR203568) in relation to the current project. JW additionally declares Stats or Methodological Editor roles for BJOG, Fertility and Sterility, Reproduction and Fertility, Journal of Hypertension, and for Cochrane Gynaecology and Fertility. AL is on the editorial board of BMC Medical Ethics. TLi declares funding from the National Eye Institute, National Institutes of Health (UG1 EY020522). TLi serves as a methodological editor for Annals of Internal Medicine, the review editor for JAMA Ophthalmology, a Co-Editor-in-Chief for Trials, and is on the editorial board for Journal of Clinical Epidemiology. WL declares funding from the Australian National Health and Medical Research Council, Medical Research Future Fund, and Ramaciotti Foundations. WL serves as an associate editor for Human Reproduction and methodological reviewer for Fertility & Sterility and F&S Reviews. SL declares funding from the University of Melbourne, Norman Beischer Medical Research Foundation, and National Health and Medical Research Council. SL is co-ordinating editor of Cochrane Gynaecology and Fertility and an editor for Human Reproduction Open, Fertility and Sterility, and Trials. FN received funding from the French National Research Agency, the French ministry of health and the French ministry of research. He is a work package leader in the OSIRIS project (Open Science to Increase Reproducibility in Science, funded by the European Union’s Horizon Europe research and innovation programme under grant agreement No. 101094725). FN is also work package leader for the doctoral network MSCA-DN SHARE-CTD (HORIZON-MSCA-2022-DN-01 101120360), funded by the European Union. TJL is an employee of The Cochrane Collaboration and interim Editor in Chief of the Cochrane Library. TA is an employee of The Cochrane Collaboration. EF is an employee of the Cochrane Collaboration and editorial board member of Cochrane Evidence Synthesis and Methods. RR is an employee of The Cochrane Collaboration and editor of Cochrane Evidence Synthesis and Methods. AA and CJV declares funding from the UKRI Metascience Unit (ESRC and Open Philanthropy) for the INSPECT-AI project. UKRI1083: Evaluating the Development and Impact of AI-Assisted Integrity Assessment of Randomised Trials in Evidence Syntheses. ES is a Senior Editor of the Cochrane Database of Systematic Reviews. LP is on the editorial board for Journal of Clinical Epidemiology. SG is an employee of the Cochrane Collaboration. SW is editor for Cochrane Anesthesia and involved in development of the Research Integrity Assessment (RIA) tool for RCTs included in evidence synthesis. ALS is on the Editorial Board for Cochrane Evidence Synthesis Methods, Statistical Editor for Cochrane Neonatal and Co-Convenor for the Cochrane Prospective Meta-Analysis Methods Group. KEH is Associate Convenor for the Cochrane Prospective Meta-Analysis Methods Group. BWM reports consultancy, travel support and research funding from Merck and consultancy for Ferring, Organon, UNILAB and Norgine. RW declares funding from the Australian National Health and Medical Research Council. RW is a Deputy Editor of Human Reproduction Update, an Editorial Board Member of BJOG and Cochrane Gynaecology and Fertility Group, and a former Deputy Editor of Human Reproduction. NOC Between 2020 and 2023 NOC was Coordinating Editor of the Cochrane Pain, Palliative and Supportive Care group, whose activities were funded by The UK National Institute of Health and Care Research (NIHR). He is the Chair of the International Association for the Study of Pain (IASP) Methodology, Evidence Synthesis and Implementation Special Interest Group and has held a grant from the ERA-NET Neuron Co-Fund. LJ is the creator of the scrutiny package in R. VB is the Editor in Chief of the Medical Journal of Australia and on the Editorial Board of Research Integrity and Peer Review. MC declares Coordinating Editor or Editor in Chief roles for James Lind Library, Cochrane Methodology Review Group and Journal of Evidence-Based Medicine, and many years of funded research using randomised trials and systematic reviews. ZA declares membership of the Cochrane Library Editorial Board, and PI on a grant from Children Investment Foundation Fund to University of Liverpool to investigate research integrity of clinical trials related to nutritional supplements in pregnancy. JDe is a member of the Cochrane Emergency and Acute Care Thematic Group. MvW is sign-off Editor for Cochrane, Senior Editor of Cochrane Gynaecology and Fertility and Cochrane Sexually Transmitted Infections, and Editor in Chief of Human Reproduction Update. JAJH declares funding from Open Philanthropy and consultancy for the Gates Foundation and Helena Foundation. HF declares an editorial role at The Lancet. All other authors declare no conflict of interest.

## Acknowledgements

The following people participated in the Delphi process and gave consent to be acknowledged: Adrian Barnett, Queensland University of Technology; Aidan G Cashin, Neuroscience Research Australia; Alessandra Alteri, IRCCS San Raffaele Scientific Institute; Amanda C de Williams, University College London; André Gillibert, University Hospital Center of Rouen; Andy Vail, University of Manchester;; Anna Abalkina, Free University of Berlin; Areti Angeliki Veroniki, Unity Health Toronto and University of Toronto; Barbara Nussbaumer-Streit, University for Continuing Education Krems; Bartosz Helfer, University of Wroclaw; Bassel H. Al Wattar, University College London; Bastiaan Van Grootven, University of Basel; Bharath Kumar Tirupakuzhi Vijayaraghavan, Apollo Hospitals; Brennan C Kahan, University College London; Changhao Liang, Beijing University of Chinese Medicine; Chris J Sutton, University of Manchester; Christina Palantza, University of Bristol; Chunhu Shi, University of Manchester; Chunli-Lu, Guangdong Pharmaceutical University; Cindy Farquhar, University of Auckland; Clara Locher, University of Rennes; David Nunan, University of Oxford; David Robert Grimes, Trinity College Dublin; David RT Laursen, Cochrane Denmark & Centre for Evidence-Based Medicine Odense (CEBMO); Dominic Ledinger, University for Continuing Education Krems; Emma Axon, Cochrane Methods Support Unit; Emma Fisher, University of Bath; Evan Mayo-Wilson, UNC Gillings School of Global Public Health; Ewelina Rogozinska, University College London; Gill Norman, University of Manchester; Gordon H Guyatt, McMaster University; Guillaume Cabanac, University of Toulouse III – Paul Sabatier; Gustav Nilsonne, Karolinska Institute; Hugh Thomas, The Lancet Gastroenterology & Hepatology; Isabelle Boutron, Université Paris Cité; Iwo Fober, University of Wroclaw; Jelena Savovic, University of Bristol; Jennifer A. Byrne, The University of Sydney; Jennifer Hilgart, Cochrane; Jo Weeks, University of Liverpool; Jo-Ana D. Chase, Cochrane; Joanne E McKenzie, Monash University; John A Loadsman, University of Sydney; Josef M Klein, unaffiliated; Julian PT Higgins, University of Bristol; Katie E Webster, University of Bristol; Katie L Stocking, University of Manchester; Kerry Dwan, Liverpool School of Tropical Medicine; Lan N Vuong, University of Medicine and Pharmacy at Ho Chi Minh City; Larissa Shamseer, Unity Health Toronto; Lesley-Anne Carter, University of Manchester; Leslie Choi, Cochrane Central Executive Team; Limbanazo Matandika, Kamuzu University of Health Sciences; Lindsay Robertson, Cochrane; Lucian Puscasiu, University of Medicine, Pharmacy, Science and Technology George Emil Palade Targu Mures; Manoj M Lalu, Ottawa Hospital Research Institute; Marek Czajkowski, Örebro University; Martin Ringsten, Cochrane Sweden; Marwah Anas El-Wegoud, Cochrane Central Editorial Service; Matthew J Page, Monash University; Michael C Ferraro, Neuroscience Research Australia; Mohamad AH Mohamad, Sohag University; Mohamed Fawzy, Ibnsina, Banon, Amshaj, Boshra IVF Centers; Mosab MR Abbas, Sohag University; Nadia Soliman, Imperial College London; Nicholas J DeVito, University of Oxford; Nicky Cullum, University of Manchester; Nipun Shrestha, University of Sydney; Norman Williams, University College London; Nurulamin M Noor, University of Cambridge; Otto Kalliokoski, University of Copenhagen; Patrick Ifeanyi Okonta, Delta State University; Paul Bramley, Sheffield Teaching Hospitals NHS Trust; Peter Bower, University of Manchester; Peter J Godolphin, University College London; Rebecka Klang, Örebro University; Reem A. Mustafa, University of Kansas Medical Center; Rickard Carlsson, Linnaeus University; Rik van Eekelen Amsterdam, University Medical Center; Rob Brierley, The Lancet Gastroenterology & Hepatology; Sarah Cotterill, University of Manchester; Simon L Turner, Monash University; Sofia Tsokani, Cochrane Methods Support Unit; Stephen JW Evans, London School of Hygiene & Tropical Medicine; Tanya Walsh, University of Manchester; Theresa HM Moore, University Hospitals Bristol and Weston NHS Foundation Trust; Tim Barker, University of Adelaide; Tim P Morris, University College London; Wai Cheng Foong, Royal College of Surgeons in Ireland and University College Dublin Malaysia Campus (RUMC); Yiqing Cai Beijing University of Chinese Medicine; Zachary Munn, University of Adelaide

The following people participated in user testing and gave consent to be acknowledged.

Angelina J. Mosley, Faculty of Homeopathy; Cecilie Jespersen, Cochrane Denmark & Centre for Evidence-Based Medicine Odense (CEBMO); Changhao Liang, Beijing University of Chinese Medicine; Christina Palantza, University of Bristol; Chunli-Lu, Guangdong Pharmaceutical University; Daniel Shaughnessy, University of Colorado Anschutz Medical Campus; Joanna Diong, University of Sydney; Flávia Geraldes, The Lancet; Jennifer Doerfler, Jena University Hospital; Jennifer Hilgart, Cochrane; Jennifer J. Ryan, Oregon Health & Science University; Jens Fust, Swedish Agency for Health Technology Assessment and Assessment of Social Services; Jose A. Calvache, Universidad del Cauca and Erasmus University MC; Lily Nicholson, University College London; Lucian Puscasiu, University of Medicine, Pharmacy, Science and Technology George Emil Palade Targu Mures; Michela Cinquini, Mario Negri Institute for Pharmacological Research; Mihaela Ivosevic, Cochrane Denmark & Centre for Evidence-Based Medicine Odense (CEBMO); Mohamed Fawzy, Ibnsina, Banon, Amshaj, Boshra IVF Centers; Petteri Sjögren, Sahlgrenska University Hospital; Ricky Turgeon, University of British Columbia; Rossella Salandra, University of Bath; Sarah Rhodes, University of Manchester; Sofia Tsokani, Cochrane Methods Support Unit; Timothy Barker, University of Adelaide; Valerie Smith, University College Dublin; Ze Freeman, King’s College London; Andrew Porter, Cancer Research UK Manchester Institute.

## Supplementary Tables

**Supplementary Table 1:**
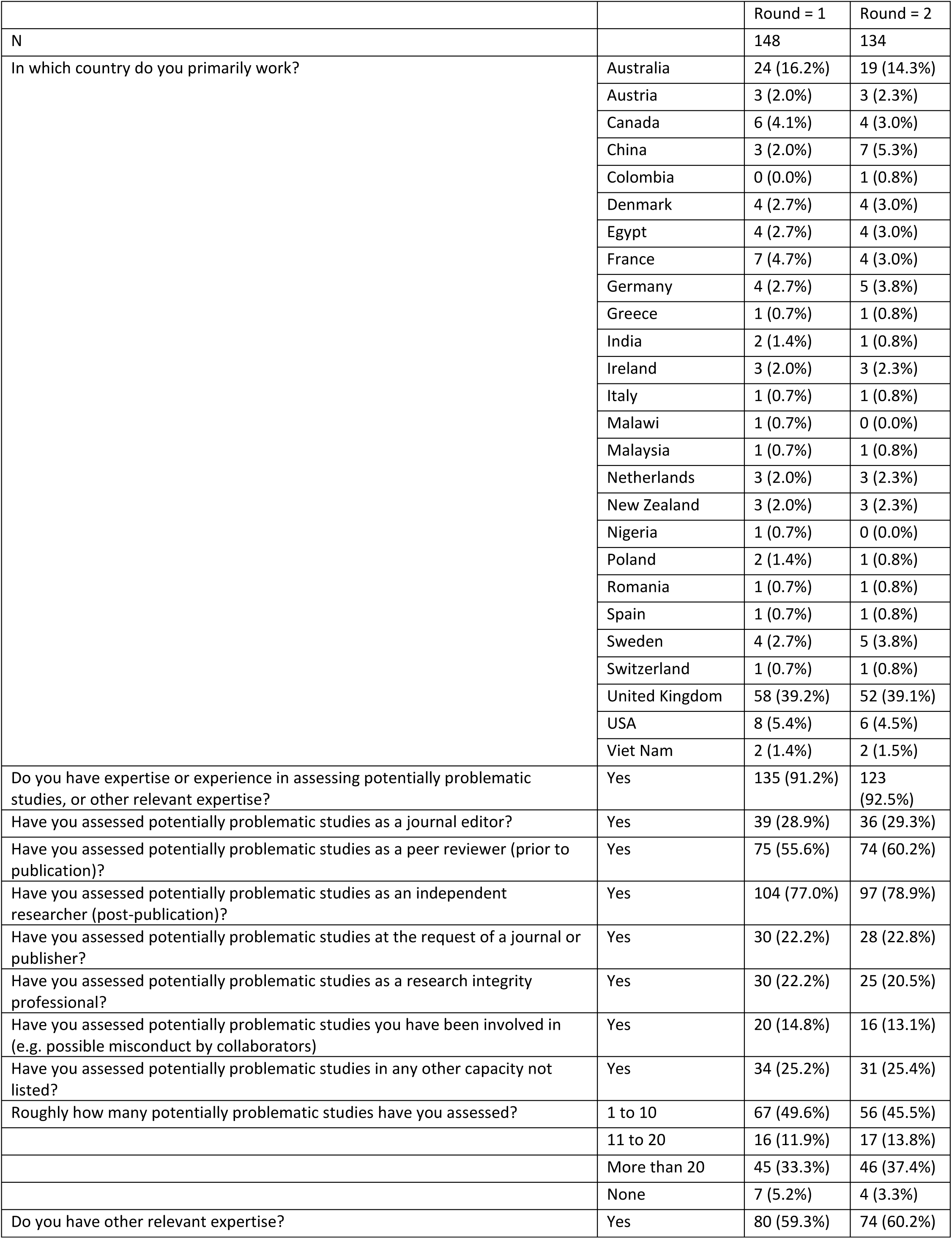
Characteristics of Delphi participants.

|  |  | Round = 1 | Round = 2 |
| --- | --- | --- | --- |
| N |  | 148 | 134 |
| In which country do you primarily work? | Australia | 24 (16.2%) | 19 (14.3%) |
|  | Austria | 3 (2.0%) | 3 (2.3%) |
|  | Canada | 6 (4.1%) | 4 (3.0%) |
|  | China | 3 (2.0%) | 7 (5.3%) |
|  | Colombia | 0 (0.0%) | 1 (0.8%) |
|  | Denmark | 4 (2.7%) | 4 (3.0%) |
|  | Egypt | 4 (2.7%) | 4 (3.0%) |
|  | France | 7 (4.7%) | 4 (3.0%) |
|  | Germany | 4 (2.7%) | 5 (3.8%) |
|  | Greece | 1 (0.7%) | 1 (0.8%) |
|  | India | 2 (1.4%) | 1 (0.8%) |
|  | Ireland | 3 (2.0%) | 3 (2.3%) |
|  | Italy | 1 (0.7%) | 1 (0.8%) |
|  | Malawi | 1 (0.7%) | 0 (0.0%) |
|  | Malaysia | 1 (0.7%) | 1 (0.8%) |
|  | Netherlands | 3 (2.0%) | 3 (2.3%) |
|  | New Zealand | 3 (2.0%) | 3 (2.3%) |
|  | Nigeria | 1 (0.7%) | 0 (0.0%) |
|  | Poland | 2 (1.4%) | 1 (0.8%) |
|  | Romania | 1 (0.7%) | 1 (0.8%) |
|  | Spain | 1 (0.7%) | 1 (0.8%) |
|  | Sweden | 4 (2.7%) | 5 (3.8%) |
|  | Switzerland | 1 (0.7%) | 1 (0.8%) |
|  | United Kingdom | 58 (39.2%) | 52 (39.1%) |
|  | USA | 8 (5.4%) | 6 (4.5%) |
|  | Viet Nam | 2 (1.4%) | 2 (1.5%) |
| Do you have expertise or experience in assessing potentially problematic studies, or other relevant expertise? | Yes | 135 (91.2%) | 123 (92.5%) |
| Have you assessed potentially problematic studies as a journal editor? | Yes | 39 (28.9%) | 36 (29.3%) |
| Have you assessed potentially problematic studies as a peer reviewer (prior to publication)? | Yes | 75 (55.6%) | 74 (60.2%) |
| Have you assessed potentially problematic studies as an independent researcher (post-publication)? | Yes | 104 (77.0%) | 97 (78.9%) |
| Have you assessed potentially problematic studies at the request of a journal or publisher? | Yes | 30 (22.2%) | 28 (22.8%) |
| Have you assessed potentially problematic studies as a research integrity professional? | Yes | 30 (22.2%) | 25 (20.5%) |
| Have you assessed potentially problematic studies you have been involved in (e.g. possible misconduct by collaborators) | Yes | 20 (14.8%) | 16 (13.1%) |
| Have you assessed potentially problematic studies in any other capacity not listed? | Yes | 34 (25.2%) | 31 (25.4%) |
| Roughly how many potentially problematic studies have you assessed? | 1 to 10 | 67 (49.6%) | 56 (45.5%) |
|  | 11 to 20 | 16 (11.9%) | 17 (13.8%) |
|  | More than 20 | 45 (33.3%) | 46 (37.4%) |
|  | None | 7 (5.2%) | 4 (3.3%) |
| Do you have other relevant expertise? | Yes | 80 (59.3%) | 74 (60.2%) |

**Supplementary Table 2:** Results of the Delphi survey. Median (IQR) shown for usefulness and feasibility scores.

| Check | Usefulness | Feasibility | % scoring 7+ usefulness |
| --- | --- | --- | --- |
| Are there typographical errors? | 4(3 to 5) | 9(8 to 9) | 12.9 |
| Has the study been retracted or does it have an expression of concern, a relevant post-publication amendment, a critical Retraction Watch or PubPeer comment or has been previously excluded from a systematic review? | 9(9 to 9) | 9(8 to 9) | 100 |
| Is there evidence of copied work, such as duplicated or partially duplicated tables or figures? | 9(8 to 9) | 5(4 to 6) | 97 |
| Is there evidence of text reuse (cutting and pasting text between papers), including text that is inconsistent with the study? | 8(7 to 8) | 6(5 to 7) | 81.8 |
| Is there evidence of automatically-generated text? | 7(5 to 8) | 5(4 to 6) | 55.8 |
| Was the study published in a predatory journal? | 7(5 to 8) | 7(6 to 8) | 61.4 |
| Is there evidence of manipulation or duplication of images? | 9(8 to 9) | 5(4 to 6) | 96.2 |
| Are important features missing from the paper? | 7(5 to 8) | 8(7 to 8) | 54.7 |
| Was the time between submission to acceptance reasonable? | 6(5 to 7) | 9(8 to 9) | 39.5 |
| Is contact information for the author team present and plausible? | 7.5(6 to 9) | 9(8 to 9) | 72.7 |
| Are any baseline data implausible with respect to magnitude, frequency, or variance? | 9(8 to 9) | 7(6 to 8) | 96.9 |
| Is the number of participants lost to follow-up compatible with the population, setting, and timeline? | 8(7 to 8) | 7(6 to 8) | 93.1 |
| Are subgroup means compatible with those for the whole cohort? | 7(7 to 8) | 7(6 to 8) | 84.7 |
| Are the reported summary data compatible with the reported range? | 8(7 to 9) | 8(7 to 9) | 93.8 |
| Are correct units reported? | 7(6 to 8) | 8(8 to 9) | 59.2 |
| Are calculations of proportions and percentages correct? | 7(6 to 8) | 9(8 to 9) | 68.5 |
| Are numbers of participants correct and consistent throughout the publication? | 8(7 to 9) | 9(8 to 9) | 82.3 |
| Are there any discrepancies between data reported in figures, tables and text? | 8(7 to 9) | 9(8 to 9) | 82.4 |
| Are any outcome data, including estimated treatment effects, implausible? | 8(8 to 9) | 7(7 to 8) | 92.4 |
| Are statistical tests correct? | 8(8 to 9) | 6(5 to 7) | 90.8 |
| Are differences in variances in baseline variables between randomised groups plausible? | 7(7 to 8) | 6(5 to 7) | 77.9 |
| Are any of the baseline data excessively **similar** between randomized groups? | 8(7 to 8) | 7(6 to 8) | 78.3 |
| Are any of the baseline data excessively **different** between randomised groups? | 7(7 to 8) | 7(6 to 8) | 75.4 |
| Are the summary outcome data identical or nearly identical across study groups? | 7(6 to 8) | 8(7 to 9) | 73.2 |
| Are there any discrepancies between the values for percentage and absolute change? | 7(6 to 8) | 7(7 to 8) | 66.7 |
| Are there any discrepancies between reported data and participant inclusion criteria? | 7(7 to 8) | 8(7 to 9) | 81.6 |
| Are the variances in biological variables surprisingly consistent over time? | 7(7 to 8) | 7(6 to 8) | 83.8 |
| Are results internally consistent? | 8.5(8 to 9) | 7(6 to 8) | 95.4 |
| Are coefficients of variation unusually similar when calculated across variables reported in the paper? | 7(6 to 7) | 7(5 to 7) | 63.2 |
| Is the amount of missing data plausible? | 7(7 to 8) | 7(6 to 8) | 76.6 |
| Are the results substantially divergent from the results of multiple other studies, e.g. in a meta-analysis? | 7(6 to 8) | 8(7 to 9) | 70.2 |
| Are terminal digits compatible with a genuine measurement process? | 7(6 to 8) | 6(5 to 7) | 64.1 |
| Are the means and variances of integer data impossible? | 9(8 to 9) | 7(6 to 8) | 94.9 |
| Is there heterogeneity across studies in degree of imbalance in baseline characteristics (in meta-analysis) | 6(5 to 7) | 6(5 to 7) | 39.4 |
| Are integer data simulated from reported summary statistics plausible? | 7(6 to 8) | 5(5 to 7) | 67.8 |
| Are the numbers of participants allocated to each group plausible given the allocation method? | 8(7 to 9) | 8(7 to 9) | 89.1 |
| Is the grant funding number identical to the number in unrelated studies? | 8(7 to 9) | 8(6 to 9) | 77.3 |
| Is a funding source reported? | 6(5 to 7) | 9(9 to 9) | 40.9 |
| Is the volume of work reported by the research group plausible, including that indicated by concurrent studies from the same group? | 7(6 to 7) | 6(5 to 7) | 56.6 |
| Is the reported staffing adequate for the study conduct as reported? | 6(5 to 7) | 5(4 to 7) | 32.5 |
| Is the recruitment and follow-up of participants plausible? | 7(7 to 8) | 7(6 to 8) | 80.6 |
| Is the interval between study completion and manuscript submission plausible? | 7(6 to 8) | 8(6 to 9) | 67.7 |
| Is there evidence that the work has been approved by a specific, recognized ethics committee? | 9(8 to 9) | 9(8 to 9) | 92.2 |
| Are there any concerns about unethical practice? | 8(7 to 9) | 7(6 to 7) | 82.9 |
| Could the study plausibly be completed as described? | 8(7 to 9) | 7(5 to 7) | 89 |
| Are the locations where the research took place specified, and is this information plausible? | 7(6 to 7) | 7(5.5 to 8) | 59.7 |
| Do the authors agree to share individual participant data, or are the data publically available? | 8(7 to 9) | 9(7 to 9) | 76.9 |
| Does the trial registration number refer to other studies? | 9(8 to 9) | 9(8 to 9) | 93 |
| Has the study been prospectively registered? | 9(7 to 9) | 9(9 to 9) | 82.2 |
| Are details such as dates and study methods in the publication consistent with those in the registration documents? | 8(7 to 9) | 9(8 to 9) | 86.2 |
| Do authors cooperate with and provide satisfactory responses to requests for information? | 8(7 to 9) | 8(6 to 9) | 82.3 |
| Is the procedure of the study aligned with local legislations? | 7(5 to 7) | 4(3 to 5) | 50.8 |
| In which country was the study conducted? | 5(3 to 6) | 9(9 to 9) | 20.2 |
| Are withdrawal and loss to follow-up rates in multiple trials by the same authors plausible? | 7(7 to 8) | 5(4 to 7) | 76.4 |
| Are contributorship statements present and complete? | 5(5 to 6) | 9(8 to 9) | 23.4 |
| Have the data been published elsewhere by the research team in an illegitimate fashion? | 9(8 to 9) | 6(5 to 7) | 90.6 |
| Are duplicate-reported **results** consistent between publications? | 8(7 to 9) | 7(5 to 8) | 88.5 |
| Are relevant **methods** consistent between publications? | 7(7 to 9) | 7(5 to 8) | 77.8 |
| Is any duplicate reporting acknowledged or explained? | 7(5 to 8) | 7.5(6 to 9) | 58.3 |
| Does the statistics methods section use generic language, suggesting lack of expert statistical input? | 6(5 to 7) | 7(6 to 7) | 31.2 |
| Are terminal digits from multiple studies by the same authors compatible with a genuine measurement process? | 7(6 to 8) | 5(4 to 6) | 57.3 |
| Does consideration of other studies from members of the research team highlight causes for concern (including expressions of concern, relevant post-publication amendment, or critical retraction)? | 8(8 to 9) | 7(7 to 8) | 95.3 |
| Are the results in multiple studies from the same author implausibly similar? | 9(8 to 9) | 6(5 to 7) | 94.5 |
| Do all authors meet criteria for authorship? | 5(4 to 6) | 5(3 to 6) | 15.3 |
| Is authorship of related papers consistent? | 5(4 to 6) | 6(4 to 7) | 14.8 |
| Are the authors on staff of institutions they list? | 6(5 to 8) | 6(5 to 7) | 44.1 |
| Do any authors have a professorial title but no other publications on PubMed? | 6(5 to 7) | 7(6 to 8) | 37 |
| Can co-authors attest to the reliability of the paper? | 7(6 to 8) | 5(4 to 6) | 63.3 |
| Given the nature of the study, does the author list make sense? | 6(5 to 7) | 6(5 to 7) | 48 |
| Are results robust to or explainable by recoding of outcomes? | 7(6 to 8) | 6(5 to 8) | 61.2 |
| Are results robust to or explainable by redaction of outcomes? | 7(5 to 8) | 6(5 to 8) | 54.1 |

**Supplementary Table 3:** Results of consensus meetings.

| Check | Meeting 1 vote to include | Meeting 2 vote to include | Consensus vote (if applicable) | Discussion points | Inclusion in tool |
| --- | --- | --- | --- | --- | --- |
| *Inspecting conduct, governance, and transparency* |  |  |  |  |  |
| Is the recruitment and follow-up of participants plausible? | 9/9 | 7/7 | NA | Recruitment and follow-up should be distinguished as separate checks. | 2.4. Is the recruitment of participants implausible?<br>4.5. Are the numbers of participants lost to follow-up implausible? |
| Is there evidence that the work has been approved by a specific, recognized committee? | 9/9 | 8/8 | NA | Should check certification of committee. Should check uniqueness of approval number and name. | 2.1. Are there concerns relating to ethical approval? |
| Are there any concerns about unethical practice? | 7/9 | 1/8 | 8/16 | Could be sign of fake study – authors have not thought through what would be acceptable. But quite vague, and potentially redundant given check of approval. | No (covered by 2.1 above) |
| Could the study plausibly be completed as described? | 7/8 | 8/8 | NA | Change wording to “conducted”. Include consideration of whether funding was sufficient. | 2.5. Are the reported methods implausible considering the reported resources? |
| Does the trial registration number refer to other studies? | 3/9 | 3/8 | NA | Can be covered by another check comparing publication details to registration documents | No (covered by 2.3 below) |
| Has the study been prospectively registered? | 9/9 | 8/8 | NA | Easy to do, so no excuse not to do it. Registering shortly after start of study might not be a cause for concern, so “prospective” might not be appropriate choice of wording. | 2.2. Are there concerns relating to the timing or absence of study registration? |
| Are details such as dates and study methods in the publication consistent with those in the registration documents? | 8/9 | 8/8 | NA | Could potentially be expanded to include comparisons with other documents (protocol, other publications) | 2.3. Are there important inconsistencies between the publication and the registration documents?<br>Comparisons with other publications covered by 4.11 |
| Do authors cooperate with requests for information? | 8/9 | 1/8 | 9/16 | May be useful flag, but need to include “appropriately”. Guidance will tell reviewers to contact authors to resolve issues; if not resolved there will be a mark against the study for the appropriate check. So no need to penalize twice. | No. Guidance states reviewers should contact authors to resolve discrepancies. |
| *Inspecting results in the paper* |  |  |  |  |  |
| Are any baseline data implausible with respect to magnitude, frequency, or variance? | 14/14 | 9/9 | NA | Could include assessment of baseline p-value distribution (that check did not meet consensus threshold in Delphi survey). | 4.3. Are any baseline data implausible?<br>Baseline p-value assessment discussed in relation to this check in guidance, but not routinely recommended. |
| Is the number of participant withdrawals compatible with the disease, age and timeline? | 12/13 | 8/8 | NA | “Withdrawals” unclear. We mean loss to follow-up for any reason. | 4.5. Are the numbers of participants lost to follow-up implausible? |
| Are numbers of participants correct and consistent throughout the publication? | 11/12 | 9/9 | NA | Participant numbers might change due to loss to follow-up or withdrawal. So need to make clear only interested in unexplained inconsistencies. | 4.6. Are there any unexplained inconsistencies in the numbers of participants? |
| Are any outcome data, including estimated treatment effects, implausible? | 14/14 | 9/9 | NA | Considered to be straightforward. | 4.7. Are any outcome data, including estimated treatment effects, implausible? |
| Are statistical test results correct? | 12/14 | 8/8 | NA | Need to consider explanations (e.g. analysis might have been adjusted for covariates.) P-value might be very different when reproduced from rounded summary data. Might be more suspicious if all results can be reproduced from rounded summary data. | 4.9. Are there errors in statistical results?<br>Guidance discusses concerns due to ability to reproduce results using summary data. |
| Are there any discrepancies between reported data and participant inclusion criteria? | 11/14 | 9/9 | NA |  | 4.1. Are there any unexplained discrepancies between reported data and participant eligibility criteria? |
| Are the variances in biological variables surprisingly consistent over time? | 1/14 | 4/9 | NA | Difficult to assess. Could potentially be considered as part of plausibility of outcome data. | No. |
| Are the means and variances of integer data impossible? | 13/14 | 5/5 | NA |  | 4.8. Are the means and variances of integer data impossible? |
| Are the numbers of participants allocated to each group plausible given the allocation method? | 12/14 | 5/8 | 14/17 | May pick up contradictions. More useful for considering all studies from one author than a single study. Don't want to overlap with risk of bias tools. Could potentially be assessed as part of plausibility of baseline characteristics? | 4.2. Are numbers of participants allocated to each group implausible given the allocation method? |
| Are any contradictions implied by the results? | Participants voted on whether to include the next four checks as sub-checks here. |  |  |  |  |
| Are subgroup means incompatible with those for the whole cohort? | 13/14 | 7/8 | NA | Considered useful, but might not frequently be testable | Included in 4.10. Are any other contradictions implied by the data? |
| Are the reported summary data compatible with the reported range? | 14/14 | 5/6 | NA |  | Included in 4.10. Are any other contradictions implied by the data? |
| Are there any discrepancies between data reported in figures, tables and text? | 10/13 | 9/9 | NA | Sufficiently important to be a standalone check | 4.4. Are there any discrepancies between results reported in figures, tables, and text? |
| Are results internally consistent? | 9/13 | 0/8 | NA | Considered unclear, even after considering examples. Would need a wording change to avoid misuse (examples of contradictory results could be indicated under the more general check) | Contradictions can be highlighted in 4.10. |
| *Inspecting text and publication details* |  |  |  |  |  |
| Has the study been retracted or does it have an expression of concern, a relevant post-publication amendment, a critical Retraction Watch or PubPeer comment or has been previously excluded from a systematic review? | 9/9 | 6/7 | NA | Spirit of the check correct but it bundles together too many distinct things. Retractions and EoC should be treated separately. Mixed views on role of PubPeer (it is possible a PubPeer comment could have no merit). Decided to include PubPeer in guidance document – reviewer might consider PubPeer comments to help them make an assessment, but presence of a comment should not automatically trigger concerns. | 1.1. Does the study have an associated retraction?<br>1.2. Does the study have an associated expression of concern or other relevant post publication notice? |
| Is there evidence of copied work, such as duplicated or partially duplicated tables? | 1/8 | 5/6 | 6/12 | Agreed that this would be useful, but there were concerns about feasibility at current time. This could be included in a check concerning plagiarism to allow instances of duplicated tables to be highlighted if noticed. Software solutions might improve feasibility. | Included in 3.1 |
| Is there evidence of text reuse (cutting and pasting text between papers), including text that is inconsistent with the study? | 8/8 | 6/7 | NA | Focus should be on inconsistent text (because plagiarism detection software not universally available). Duplicated text can be highlighted with this check if detected. | 3.1. Are there concerns relating to duplicated content, such as text or tables, or text that is incompatible with the study? |
| Is there evidence of manipulation or duplication of images? | 7/7 | 5/7 | NA | Unclear whether software is useful for the types of figures found in RCT publications. Manual checks useful in the absence of software. | 3.2. Is there evidence of manipulation or duplication of figures? |
| *Inspecting the research team and their other work* |  |  |  |  |  |
| Have the data been published elsewhere by the research team in an illegitimate fashion? | 1/8 | 4/12 | NA | Not relevant to trustworthiness of the RCT. | No |
| Are duplicate-reported data consistent between publications? | 8/8 | 8/9 | NA | Consistency of methods is just as important as consistency of results, so should be added. | 4.11. Are there inconsistencies in descriptions of methods and results across publications describing the study? |
| Does consideration of other studies from members of the research team highlight causes for concern (including expressions of concern, relevant post-publication amendment, or critical retraction)? | 8/8 | 11/11 | NA | Discussion about whether track record of the author team should be used to assess the index RCT. | 1.3. Do other studies by the research team highlight causes for concern (associated retractions, expressions of concern, relevant post-publication notices?) |
| Are the results in multiple studies from the same author implausibly similar? | 1/8 | 3/12 | NA | Not feasible to consider all studies from the author team. | No |

**Supplementary Table 4:**
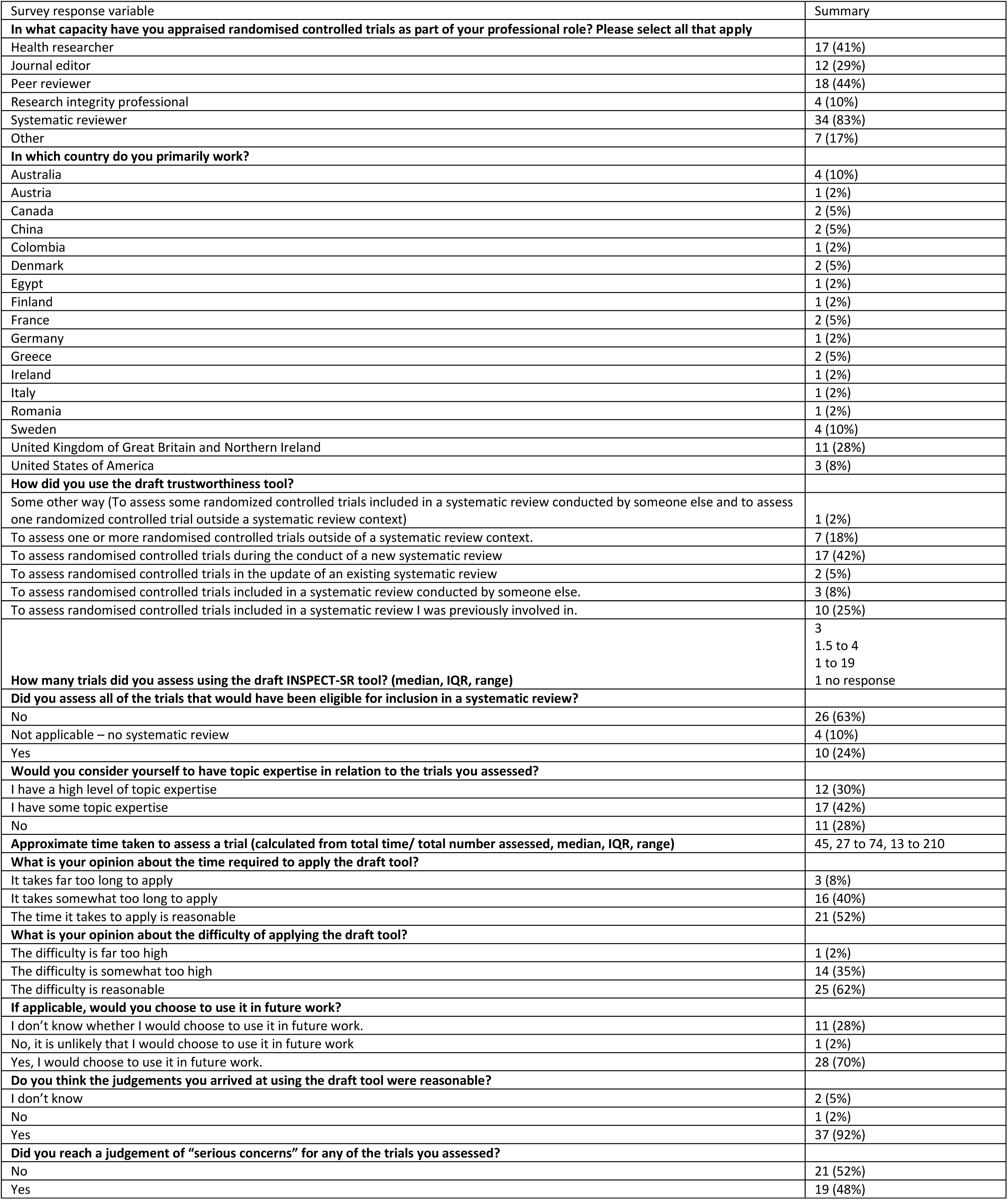
Summary of quantitative responses to user testing survey.

**Supplementary Table 5:** Anonymised minutes from user testing workshop.

| Topic | Summary of discussion |
| --- | --- |
| Structure of the tool | Liked how it was similar to ROBIS, felt familiar, not overwhelming<br>(People generally happy with this) |
| Approach to arriving at domain-level judgements | Even if 'some concerns' across all checks, wouldn't have moved domain to judgement to 'severe concerns'<br>+1 agree<br>Needs to be done by two people, preferably independently – people must have domain and stats proficiency (between them)<br>Prefer not algorithm driven (1 person)<br>Full guidance doc will help a lot<br>If not able to assess (due to lack of knowledge), what to use – 'unclear'? JW confirmed 'unclear' to be used that way<br>More difficult than risk of bias, more reliant on individual knowledge/communication with review authors<br>When systematic reviews are published would these INSPECT results be included? – JW: yes, articulation of judgement should be clear<br>JW: Should 'no information' be a domain level option? [e.g. if all checks in the domain unclear]<br>+1 against this<br>+1 for this<br>Need guidance on contacting authors to be consistent with Cochrane policy |
| Approach to arriving at an overall study-level judgement | JW: Should no prospective registration warrant 'serious concerns'?<br>+3 for no (expectations vary by region too greatly)<br>1 felt legality should be cut-off for expectation of registration<br>No other issues here |
| Time to complete | I don't see how it could be shorter, thinks 60-75 minutes wouldn't be too long, given the stakes. Was easier with studies I was familiar with<br>"Are there errors in statistical results" can be extremely time consuming. Resources to help people with that would be v.useful.<br>GRIM/GRIMMER v.useful and didn't take long<br>Papers that took longest were those that had 'some' concerns over multiple domains, required more in-depth investigation to determine final judgement.<br>Maybe these can be done as you're going along (i.e. you'll be reading the paper anyway as part of the systematic review, just do it then).<br>Order checks by severity (those most likely to find problems)? JW: We've had a go at doing that<br>JW: can potentially save time doing ROB if do this first |
| Difficulty | automation: awarded grant to help automate some of these checks |
|  | Cochrane should have some central plagiarism check, should maybe be up to them<br>Duplicated text domain – if no information [no plagiarism software, no figures] the domain doesn't contribute anything (and therefore the weights of the other domains increase)<br>Plausibility (of results) checks – a sensitive subject, worried any area where mechanism of action is unknown may be targeted by reviewer with nefarious motivation (complementary med). Maybe get Cochrane Complementary to have a look<br>Plausibility is not just effect or not, about magnitude also. Reiterated point that having 2<sup>nd</sup> rater will help with this issue<br>Plausibility of other things (recruitment, etc), were helpful/uncontroversial. Guidance on this can make a big difference to ability to judge these checks<br>JW: Okay to update checks/ROB in light of meta-analysis? (i.e. noticing substantial difference in effect size in forest plot)<br>+2 for yes<br>+1 for no – should be fresh process every time, can't be any post-hoc retrospective work<br>Maybe a technical/reporting issue – what do you report, how to ensure transparency if you do go back and change assessment? |
| Any particularly challenging domains or checks? | JW: Open Q for 'other' concerns?<br>Already some free text boxes that can serve this purpose [JW: people put rubbish into 'other' boxes, and we don't want people using checks we don't endorse].<br>JW: reasonable to allow people to do their own order of domains?<br>First check must come first, otherwise – probably doesn't matter<br>Identification of 'critical' domain/checks may be option (as in AMSTART/ROBIS) |
| Overall impression, suggestions | JW: is scope of tool clear?<br>+1 for yes<br>Is application of tool going to be web-form or downloadable word doc?<br>JW: specifics of implementation to be worked out |
| Other | Definition of problematic studies (from Cochrane) includes unpublished study. Perhaps INSPECT should also? [JW: think it could be used, probably some domains unclear]<br>Should be more pointed mention of pubpeer plugin.<br>Problematic paper screener website? (for tortured phrases) |

